# The Role of Disease Severity and Demographics in the Clinical Course of COVID-19 Patients Treated with Convalescent Plasma

**DOI:** 10.1101/2021.01.19.21249678

**Authors:** Tengfei Ma, Chad C. Wiggins, Breanna M. Kornatowski, Ra’ed S. Hailat, Andrew C. Clayburn, Winston Guo, Patrick W. Johnson, Jonathon W. Senefeld, Stephen A. Klassen, Sarah E. Baker, Katelyn A. Bruno, DeLisa Fairweather, R. Scott Wright, Rickey E. Carter, Chenxi Li, Michael J. Joyner, Nigel Paneth

**Affiliations:** Department of Epidemiology and Biostatistics, College of Human Medicine, Michigan State University, East Lansing, Michigan; Department of Anesthesiology and Perioperative Medicine, Mayo Clinic, Rochester, Minnesota; Department of Health Sciences Research, Mayo Clinic, Jacksonville, Florida; Department of Cardiovascular Medicine, Mayo Clinic, Jacksonville, Florida; Department of Cardiovascular Medicine and Director Human Research Protection Program, Mayo Clinic, Rochester, Minnesota; Department of Pediatrics and Human Development, College of Human Medicine, Michigan State University, East Lansing, Michigan

## Abstract

Treatment of patients with COVID-19 using convalescent plasma from recently recovered patients has been shown to be safe, but the time course of change in clinical status following plasma transfusion in relation to baseline disease severity has not yet been described. We analyzed short, descriptive daily reports of patient status in 7,180 hospitalized recipients of COVID-19 convalescent plasma in the Mayo Clinic Expanded Access Program. We assessed, from the day following transfusion, whether the patient was categorized by his or her physician as better, worse or unchanged compared to the day before, and whether, on the reporting day, the patient received mechanical ventilation, was in the ICU, had died or had been discharged. Most patients improved following transfusion, but clinical improvement was most notable in mild to moderately ill patients. Patients classified as severely ill upon enrollment improved, but not as rapidly, while patients classified as critically ill/end-stage and patients on ventilators showed worsening of disease status even after treatment with convalescent plasma. Patients age 80 and over showed little or no clinical improvement following transfusion. Clinical status at enrollment and age appear to be the primary factors in determining the therapeutic effectiveness of COVID-19 convalescent plasma among hospitalized patients.

---

The number of deaths from COVID-19 in the United States surpassed 350,000 on January 3, 2021^1^, just ten months after the first confirmed case of novel coronavirus (SARS-CoV-2) in the US, demonstrating the urgent need to find safe and effective treatment options. Convalescent plasma, rich in antibodies from recently recovered patients, was used successfully in the 1918 influenza pandemic^2^, SARS-1^3^ and Ebola^4^ epidemics. Recognizing that a vaccine would not be widely available for several months to a year, and facing a paucity of treatment options, the United States Federal Government, in collaboration with the Mayo Clinic and the national blood banking community, developed the Expanded Access Program (EAP) for COVID-19 convalescent plasma as a national registry to examine the safety and as much as possible the efficacy of convalescent plasma treatment in hospitalized patients. The inclusion criteria of the EAP required that enrolled patients 1) have a diagnosis of SARS-CoV-2, 2) be severely ill or at high risk for becoming severely ill from COVID-19, and 3) be admitted to an acute care facility for COVID-19 complications.

Evidence of efficacy has emerged from retrospective comparisons of treated and untreated patients^4^, and from several small randomized trials^5–12^. However, only one of these trials^13^ and one retrospective treatment-control analysis^14^ in hospitalized patients stratified patients based on disease severity at time of treatment to examine efficacy accordingly. Analyses of the EAP have shown that COVID-19 convalescent plasma is safe^15,16^ and likely to be effective in treating COVID-19^17^. Based on the historical literature, we hypothesized that patients treated with convalescent plasma earlier in the course of the disease (who were not on mechanical ventilation or in the intensive care unit (ICU)) or who had less severe disease at the time of transfusion, would show more rapid and better improvement than convalescent plasma recipients receiving mechanical ventilation or in the ICU at the time of treatment.

## THE RAPID EVALUATION PROJECT (REP)

The EAP was developed primarily as a registry to investigate the safety of convalescent plasma as a treatment for COVID-19 during an ongoing pandemic. It was implemented at a time when health systems were overwhelmed and clinical research resources were limited because of hospitals restricting patient access to essential personnel and the frequent reassignment of research staff to clinical duties. We therefore developed the REP as an optional reporting tool requiring minimal time and effort on the part of the treating physician, but that would nonetheless provide useful information as to whether improvement or worsening was noted following treatment with convalescent plasma, and how this varied by category of patient.

We offered all physicians enrolling patients in the EAP the opportunity to participate in the Rapid Evaluation Project (REP) by providing brief, daily updates of the status of their patients until death or discharge from the hospital. Physicians or their designees, after providing the baseline status of each participant on the day of enrollment, described each participant’s status compared to the previous day – better, the same, or worse - and also noted whether the patient had been discharged from hospital or died that day, and whether the patient was in an ICU or had required mechanical ventilation.

This simple system was inspired by the study by Waller and Lawther who asked London patients with chronic obstructive pulmonary disease to record their daily status – ‘better, worse, much worse, or the same as usual’ - and used these scores to document a clear relationship of worse days to specific components of air pollution^18,19^.

## RESULTS

### Patient Demographics and Baseline Severity

Sufficient follow-up information was submitted on 8,311 convalescent plasma recipients. Descriptions of the demographics and disease characteristics are presented in Table 1. 534 patients were excluded due to receipt of multiple transfusions during their hospitalization, and another 597 patients who had less than two days of follow-up data, leaving a total of 7,180 patients for analysis. Participants in the REP were very similar to the overall EAP population, at the time of data analysis (August 1, 2020), in age distribution, sex, race and initial clinical status, but tended to have slightly more respiratory risk factors such as dyspnea, low oxygen parameters and extensive early lung infiltrates. Compared to the entire EAP cohort (Table 1, third column), there were fewer Hispanic/Latino patients and more patients residing the Midwest and the West regions of the U.S, and fewer in the Northeast. Patients were severely ill, with 53% in an ICU and 28% requiring mechanical ventilation prior to transfusion with convalescent plasma.

**Table 1.** Demographic factors and disease severity in patients in the Rapid Evaluation Program and the overall Expanded Access Program^†^ patients.

|  | Rapid Evaluation<br>Project Sample<sup>†</sup> | Total EAP<br>Enrollment<sup>†</sup> |
| --- | --- | --- |
| **Patient Characteristics** |  |  |
| Total patients enrolled | 9,752 | 50,385 |
| Enrolled patient received a transfusion | 8,311 | -- |
| Patients excluded due to insufficient follow-up<sup>a</sup> | 381 | -- |
| Received >1 transfusion<sup>b</sup> | 534 | 2,522 (5.0%) |
| Patients included in the analysis | 7,180 (100%)* | 50,385 (100%)* |
| **Patient Outcomes** |  |  |
| Discharge reported<sup>c</sup> | 4,435 (60.0%) | 26,710 (53.0%) |
| Death reported | 1,659 (22.4%) | 10,947 (21.7%) |
| **Geographic Region** |  |  |
| Midwest | 1,521 (20.6%) | 8,046 (16.0%) |
| Northeast | 949 (12.8%) | 10,650 (21.1%) |
| Puerto Rico | 13 (0.2%) | 47 (0.1%) |
| Southeast | 2,319 (31.4%) | 14,737 (29.3%) |
| Southwest | 1,107 (15.0%) | 9,919 (19.7%) |
| West | 1,487 (20.1%) | 6,983 (13.9%) |
| **Categorical Age (yr)** |  |  |
| 18 to 34 | 418 (5.7%) | 2,813 (5.6%) |
| 35 to 54 | 2,043 (27.6%) | 13,348 (26.5%) |
| 55 to 79 | 4,085 (55.2%) | 28,479 (56.5%) |
| 80 or older | 850 (11.5%) | 5,745 (11.4%) |
| **Gender** |  |  |
| Female | 2,974 (40.3%) | 20,280 (40.4%) |
| Male | 4,395 (59.6%) | 29,931 (59.6%) |
| Undisclosed | 8 (0.1%) | 43 (0.1%) |
| **Race** |  |  |
| Asian | 261 (3.5%) | 1,820 (3.6%) |
| Black or African American | 1,464 (19.8%) | 9,517 (18.9%) |
| Other or Unknown | 1,903 (25.7%) | 12,706 (25.2%) |
| White | 3,768 (50.9%) | 26,342 (52.3%) |
| **Ethnicity** |  |  |
| Hispanic/Latino | 2,585 (35.0%) | 19,200 (38.1%) |
| Not Hispanic/Latino | 4,811 (65.0%) | 31,185 (61.9%) |
| **Initial Clinical Status** |  |  |
| Mild or Moderate | 2,387 (34.2%) | -- |
| Severe | 3,004 (43.1%) | -- |
| Critical or End Stage | 1,565 (22.5%) | -- |

<sup>†</sup> All enrollment information as of August 3, 2020
\* Following subsections are based on this value
<sup>a</sup> Insufficient follow-up include, no daily reporting for a patient, no dates of submitted reports
<sup>b</sup> Defined as >4 h between transfusions. Multiple units given within 4 h would be considered a single transfusion
<sup>c</sup> Number of patients reported discharged or expired within 21 days of convalescent plasma treatment
<sup>d</sup> These data include a subset of the sample (n = 32,928) - only those patients that currently have severe or life-threatening COVID-19.

### Score Trajectories Following Transfusion

We constructed ordinal scales of clinical outcome from the physicians’ reports ranging from −2 to +2. Death was scored as −2, clinical worsening as −1, no change as 0, clinical improvement as +1, and hospital discharge as +2. Figure 1 displays the trajectories of mean ordinal scale scores for all patients (Figure 1A) and for several patient sub-groups (Figures 1B-1G) for the first 21 days following CP transfusion, with gray bands surrounding score trajectories indicating the 95% confidence intervals. More detailed methods regarding the daily patient scoring can be found in the data analysis section.

**Figure 1.**
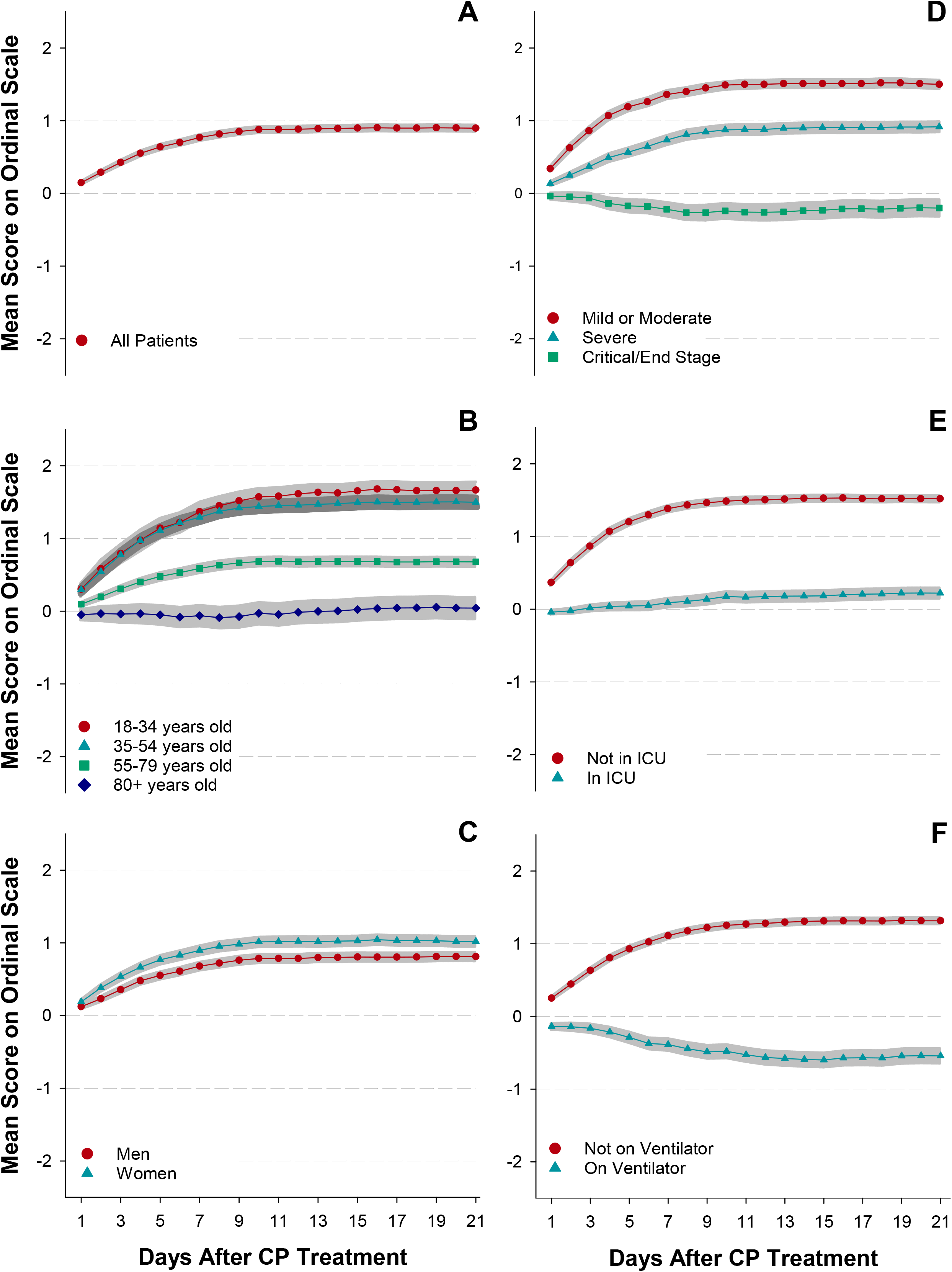
Trajectories of Daily Status Improvement/Worsening. Subgroup analyses include A) all patients, B) by age, C) by sex, D) by initial clinical status, E) by ICU status prior to transfusion, and F) by ventilator status prior to transfusion. Net patient scores of 0= no net change, +1= net improvement but still hospitalized, −1= net worsened and still hospitalized, +2= discharged from hospital, and −2= patient expiration. Points are the mean score for each day, and gray bands indicate the 95% confidence interval for each data set.

For all patients in the REP (Figure 1A), the mean daily score increased rapidly from one day after CP infusion (mean=0.16, 95% CI: 0.14 - 0.19) to day 8 (mean=0.82, 95% CI:0.78 - 0.87). Thereafter, the mean scores stabilized between 0.82 to 0.91.

When stratified by age group (Figure 1B), only the elderly (80+ years old) did not improve following transfusion but remained on average in the same status as at the time of transfusion two weeks following infusion (Day 14, mean= 0.002, 95% CI: −0.15, 0.15). While patients under 55 years of age improved more rapidly than patients 80 years and older, no substantial difference in improvement was seen between patients aged 18-34 years and patients aged 35-54. Overall, women tended to improve slightly more rapidly than did men, but both sexes exhibited overall improvement following treatment with convalescent plasma (Figure 1C).

When stratified by baseline category of illness severity (Figure 1D), patients with mild/moderate illness and severe illness improved most rapidly through day 7 and 9, respectively, and then leveled off. Patients described as critical/end-stage worsened from day 1 (mean= −0.04, 95% CI: −0.15, −0.04) onward following the transfusion to day 9 (mean= −0.26, 95% CI: −0.37, −0.16) and then slightly improved but with scores still remaining negative from day 9 to day 21 (mean= −0.19, 95% CI: −0.31, −0.08). Among all patients discharged from the hospital during the period of observation, the median length of stay following transfusion was 6 days (IQR= 4-11 days), and among patients who died, median length of stay following transfusion was 8 days (IQR= 4-13 days).

The leveling off of improvement after day 9 in the overall patient population (Figure 1A) appears to be attributable to patients still being hospitalized after that time may remain critically ill but medically stable. Very long lengths of stay have been observed in some patients surviving with COVID-19^20^. Patients not in the ICU at the time of convalescent plasma treatment (Figure 1E) improved more steadily than patients in the ICU. Patients who did not require mechanical ventilation at the time of convalescent plasma treatment (Figure 1F) had better improvement than patients requiring mechanical ventilation. Patients on ventilators declined steadily for two weeks, after convalescent plasma treatment (mean= −0.59, 95% CI: −0.69, −0.50).

Our analysis revealed that the trajectories of improvement or worsening change over time, and our statistical program was able to detect the break points indicating where the curve changed, which in all cases was a levelling off of the initial trajectory. Table 2 shows the day of the segment breakpoint for several sub-groups of the study population. The largest factor determining both direction of the trajectory and the rapidity of stabilization was ventilator status, with unventilated patients stabilizing, after improvement, after 7-8 days, while ventilated patients did not begin to stabilize from their downhill course until 12-13 days.

**TABLE 2:** Segment breakpoints in first segment coefficient by patient characteristics

| Subgroup Category | Segment Breakpoint | First Segment Coefficient |
| --- | --- | --- |
| **Overall Cohort** | 7.4 | 0.102* |
| **Age** |  |  |
| 18-34 years old | 7.6 | 0.171* |
| 35-54 years old | 6.5 | 0.183* |
| 55-79 years old | 7.5 | 0.081* |
| 80+ years old | 7.8 | -0.007 |
| **Sex** |  |  |
| Males | 7.5 | 0.092* |
| Females | 7.3 | 0.115* |
| **Disease Severity** |  |  |
| Mild or moderate | 6.4 | 0.186* |
| Severe | 8.3 | 0.096* |
| Critical or end stage | 8.4 | -0.034* |
| **ICU Status** |  |  |
| Not in ICU | 6.4 | 0.185* |
| In ICU | 10.4 | 0.021* |
| **Ventilator Status** |  |  |
| Not on ventilator | 7.4 | 0.143* |
| On ventilator | 12.3 | -0.043* |

All regression models were analyzed with 2 segments to assess changes and trends of the clinical outcome of each day.
\* indicates $P < 0.001$ for that segment of the trajectory.

**Table 3:** Multivariate analysis of improvement trajectory based on demographics and disease severity prior to treatment with COVID-19 convalescent plasma

| Variables | OR (95% CI) | P value |
| --- | --- | --- |
| **Intensive Care Status** |  |  |
| ICU admission prior to transfusion | 0.46 (0.42-0.51) | <0.0001 |
| **Mechanical Ventilation Status** |  |  |
| On ventilator prior to transfusion | 0.42 (0.37-0.47) | <0.0001 |
| **Sex** |  |  |
| Females | 1.00 (Ref) | -- |
| Males | 0.93 (0.86-1.01) | 0.070 |
| **Age** |  |  |
| 18-34 years | 1.00 (Ref) | -- |
| 35-54 years | 0.85 (0.73-0.99) | 0.044 |
| 55-79 years | 0.50 (0.43-0.58) | <0.0001 |
| 80+ years | 0.26 (0.22-0.31) | <0.0001 |
| **Odds ratio of Net Improvement by Day 7 Stratified by Disease Severity** |  |  |
| Mild or moderate | 1.00 (Ref) | -- |
| Severe | 0.45 (0.39-0.52) | <0.0001 |
| Critical or end stage | 0.21 (0.17-0.25) | <0.0001 |
| **Odds ratio of Net Improvement by Day 14 Stratified by Disease Severity** |  |  |
| Mild or moderate | 1.00 (Ref) | -- |
| Severe | 0.57 (0.45-0.73) | <0.0001 |
| Critical or end stage | 0.31 (0.23-0.41) | <0.0001 |
| **Odds ratio of Net Improvement by Day 21 Stratified by Disease Severity** |  |  |
| Mild or moderate | 1.00 (Ref) | -- |
| Severe | 0.65 (0.44-0.98) | 0.039 |
| Critical or end stage | 0.60 (0.39-0.92) | 0.019 |

Adjusted GEE model was used to estimate the odds of being in a higher category (net improvement) of the ordinal clinical outcomes.
\*Initial status of severity and time interaction term were added in the model as time-varying covariate.

### Multivariate Analysis of Factors Associated with Trajectories

To assess the individual and combined contributions of the factors described above (age, gender, ICU status, ventilator status at the time of transfusion, and a qualitative measure of baseline illness severity), we undertook a multivariate approach (Table 2), using a generalized estimating equations (GEE) model. The cumulative odds ratios from the GEE model indicate the odds for being in a higher category of the ordinal scale (net improvement or +1 on ordinal scale compared to day of infusion). Advanced age (≥80 years old) (OR = 0.26, 95% CI, 0.22-0.31, *P* <0.0001), and critical illness upon enrollment were the strongest predictors of non-improvement following convalescent plasma treatment. ICU (OR = 0.46, 95% CI, 0.42-0.51, *P* <0.0001) and ventilator status (OR = 0.42, 95% CI, 0.37-0.47, *P* <0.0001) were next in importance, and gender (OR = 0.93, 95% CI, 0.86-1.01, *P* =0.07) showed little discrimination once other factors were taken into account.

By day 7, patients who were critical/end stage were 80% less likely to show improvement compared to mild/moderate cases (OR= 0.21, 95% CI, 0.17-0.25, *P* <0.001), but by day 21, critical/end stage patients were just 40% less likely to have improved than the most favorable group (OR= 0.60, 95% CI, 0.39-0.92, *P* =0.019). The severe illness group showed substantially less improvement than the mild/moderate group at both day 7 (OR= 0.45, 95% CI, 0.39-0.52, *P* <0.0001) and day 21 (OR= 0.65, 95% CI, 0.44-0.98, *P* =0.039), although as with critically ill patients, the odds ratio of the severely ill did show some signs of improvement over time.

## DISCUSSION

The aim of the Rapid Evaluation Project was to answer whether, how quickly, and to what extent COVID-19 patients improved following the transfusion of convalescent plasma. Our data describes what happens after convalescent plasma is given in different sub-groups of patients, and may provide guidance for physicians about the best candidates to receive convalescent plasma to treat COVID-19. We cannot, of course, conclude that these trajectories are solely the result of convalescent plasma infusion.

We found that most patients who received COVID-19 convalescent plasma improved within the first 7-10 days following the transfusion. The trajectory of change in patients infused with convalescent plasma varied greatly depending on their disease state at the onset of treatment, and we found that some categories of patients did not appear to show any clinical benefit from convalescent plasma treatment during the first 21 days after treatment, including a) patients who were critical or end stage at the time of enrollment, b) patients over the age of 80, and c) those on a ventilator at the time of treatment. The lattermost group actually showed clinical deterioration following transfusion. Our results also suggest that the rate of clinical improvement in the overall cohort starts to slow down from day 7, and more noticeably after day 9 from treatment with convalescent plasma. Despite the slowing of clinical improvement after day 9, in patients not critically ill, some modest improvement continued at least until 21 days. Stability of status after the first week of treatment could also mean that convalescent plasma offers a buffer in time to allow the immune system to mount its own response to the infection and might mitigate the need for later ICU admission or mechanical ventilation. At the same time, the slowing of the rate of improvement might suggest that a second dose of passive antibody could improve outcomes if given 7-10 days after the first dose in patients whose improvement was sub-optimal^14^.

Our analysis provides an estimate of effect size in risk categories while controlling for other risk factors. We found that several factors ascertained at the time of transfusion were associated with condition change following convalescent plasma treatment including patients’ level of severity; ventilator use, ICU presence, and advanced age. We also observed that baseline severity is also related to the slope of the change in trajectory, suggesting that critically ill patients do improve following treatment with convalescent plasma, albeit at a much slower rate.

Overall, the data from our analysis provide a framework for best case use when considering administering convalescent plasma to treat COVID-19. These data offer a qualitative supplement to more quantitative analyses from our group^17^ supporting the conclusion that convalescent plasma is a valuable treatment option for many patients hospitalized due to complications from COVID-19 infection, but most especially if provided early in the course of disease before patients require ventilation or are admitted to the ICU.

## METHODS

### Study Design

The Mayo Clinic Expanded Access Program (EAP) was a national, multicenter, open-label registry of hospitalized adults with severe/life-threatening or at high risk for becoming severe COVID-19 disease, whose physicians treated, or planned to treat them with convalescent plasma. The initiation and approval of the program have been described in detail.^5,6^ Briefly, hospital and physician registration occurred through the EAP central website, www.uscovidplasma.org. The web-based registration, compliance, and data entry system for the EAP went live on April 3, 2020, and the first transfusion was given on April 7, 2020. Written informed consent was obtained from the patient or legally-authorized representative prior to enrollment, except for those patients who used an emergency consent process defined in collaboration with the United States Food and Drug Administration (21 CFR 50.23). The study was approved and overseen by the Mayo Clinic Institutional Review Board (IRB #20-003312).

The Rapid Evaluation Project (REP) was an optional sub-study of the EAP. The REP survey was implemented on May 5, 2020. Physicians who opted to participate were asked to answer three simple questions each day regarding the patient’s status compared to the day before. The survey asked 1) has the patient been in the ICU in the past 24 hours? 2) has the patient required mechanical ventilation in the past 24 hours?, and 3) how has the patient’s status changed in the last 24 hours?. Possible answers to question 3 were: a) patient was discharged from hospital, b) patient improved c) patient stayed the same, d) patient worsened, or e) patient died. Physicians and/or their designee received an automatic daily notification by email requesting the status update.

Participant enrollment in the REP was open to any patient (patient inclusion/exclusion criteria for EAP described previously^15,16^) in the EAP whose physician or designee were willing to provide daily updates.

### Data Analysis

To define each patient’s overall level of improvement or worsening on each day of observation, we calculated the total number of “condition worsened” and “condition improved” responses for each patient for the days preceding the day of observation. If a participant had more “condition worsened” than “condition improved” responses, but were still alive and still hospitalized, we defined the overall clinical outcome as clinical worsening (−1) for that day. If the participant had a greater number of “condition improved” responses, while still hospitalized, we defined the overall clinical outcome as clinical improvement (+1) for that day. If the participant had the same total number of “condition worsened” and “condition improved” responses, but still hospitalized, we defined the overall clinical outcome as no change (0). Finally, death and discharge were also considered clinical outcomes giving us five potential clinical outcomes to include in our scale. We used a 5-point ordinal scale to quantify these clinical outcomes as follows: −2 points, death; −1 point, clinical worsening; 0 points, no change; 1 point, clinical improvement; 2 points, discharge. To plot group trajectories, we averaged the ordinal scales of the patient group for each day of observation. To account for the status of all patients, including those discharged or died, patients were scored as either +2 or −2 on each day from their discharge or death until day 21.

To more fully assess the trajectories, we performed a linear segmented analysis to identify the changes of scores overtime in each patient group^21–23^. This method allowed us to test for significantly increasing or decreasing linear trends in clinical outcomes after transfusion.

The program examined the data to identify one breakpoint for each segmented regression model in which the trajectory changed The R package “segmented” was used for the data analysis[citation 19 20] and P < 0.001 was considered statistically significant.

### Multivariate Analysis

A generalized estimating equations (GEE) approach with a logit link and independence “working” correlation structure was used to study the effect of age, sex, level of severity, baseline ventilator use, and baseline ICU admission when the transfusion was administered. The model included the clinical status (worsening/improvement) defined above on days 7, 14 and 21 as repeated outcome measures. Patients who died or were discharged on days other than day 7, 14, or 21 were considered deceased or discharged on the next interval. For example, a patient who died on day 8 would be categorized as deceased on day 14. To model the effect of baseline level of severity on different time points, the regression model included a time-varying term for baseline level of severity. All statistical analyses were completed using R version 4.0.2. R package “multgee” was used for GEE analysis of ordinal multinomial responses^24,25^ and P < 0.05 was considered statistically significant.

## Data Availability

The dataset for this study is not currently publicly available.

## Notes

**Funding:** This project has been funded in part with Federal funds from the Department of Health and Human Services; Office of the Assistant Secretary for Preparedness and Response; Biomedical Advanced Research and Development Authority under Contract No. 75A50120C00096. Additionally, this study was supported in part by National Center for Advancing Translational Sciences (NCATS) grant UL1TR002377, National Institute of Diabetes and Digestive and Kidney Diseases (NIDDK) 5T32DK07352 (to CCW), National Heart, Lung, and Blood Institute (NHLBI) grant 5R35HL139854 (to MJJ) and grant 1F32HL154320 (to JWS), Natural Sciences and Engineering Research Council of Canada (NSERC) PDF-532926-2019 (to SAK), National Institute of Allergy and Infectious Disease (NIAID) grants R21 AI145356, R21AI152318 and R21 AI154927 (to DF), Schwab Charitable Fund (Eric E Schmidt, Wendy Schmidt donors), United Health Group, National Basketball Association (NBA), Millennium Pharmaceuticals, Octapharma USA, Inc, and the Mayo Clinic.

### Competing Interest Statement

The authors have declared no competing interest.

### Clinical Trial

NCT04338360

### Funding Statement

This project has been funded in part with Federal funds from the Department of Health and Human Services; Office of the Assistant Secretary for Preparedness and Response; Biomedical Advanced Research and Development Authority under Contract No. 75A50120C00096. Additionally, this study was supported in part by National Center for Advancing Translational Sciences (NCATS) grant UL1TR002377, National Institute of Diabetes and Digestive and Kidney Diseases (NIDDK) 5T32DK07352 (to CCW), National Heart, Lung, and Blood Institute (NHLBI) grant 5R35HL139854 (to MJJ) and grant 1F32HL154320 (to JWS), Natural Sciences and Engineering Research Council of Canada (NSERC) PDF-532926-2019 (to SAK), National Institute of Allergy and Infectious Disease (NIAID) grants R21 AI145356, R21AI152318 and R21 AI154927 (to DF), Schwab Charitable Fund (Eric E Schmidt, Wendy Schmidt donors), United Health Group, National Basketball Association (NBA), Millennium Pharmaceuticals, Octapharma USA, Inc, and the Mayo Clinic.

### Author Declarations

The study was approved and overseen by the Mayo Clinic Institutional Review Board (IRB #20-003312)

## References

1. U.S. Department of Health & Human Services. CDC COVID Data Tracker. (2020).

2. Luke, T. C., Kilbane, E. M., Jackson, J. L. & Hoffman, S. L. Meta-analysis: convalescent blood products for Spanish influenza pneumonia: a future H5N1 treatment? Ann. Intern. Med. 145, 599–609 (2006).

3. Mair-Jenkins, J. et al. The effectiveness of convalescent plasma and hyperimmune immunoglobulin for the treatment of severe acute respiratory infections of viral etiology: a systematic review and exploratory meta-analysis. J. Infect. Dis. 211, 80–90 (2015).

4. Van Griensven, J. et al. Evaluation of convalescent plasma for Ebola virus disease in Guinea. N. Engl. J. Med. 374, 33–42 (2016).

5. Li, L. et al. Effect of Convalescent Plasma Therapy on Time to Clinical Improvement in Patients With Severe and Life-threatening COVID-19: A Randomized Clinical Trial. J. Am. Med. Assoc. (2020).

6. Gharbharan, A. et al. Convalescent Plasma for COVID-19. A randomized clinical trial. medRxiv (2020) doi:10.1101/2020.07.01.20139857.

7. Avendano-Sola, C. et al. Convalescent Plasma for COVID-19: A multicenter, randomized clinical trial. medRxiv 2020.08.26.20182444 (2020) doi:10.1101/2020.08.26.20182444.

8. Rasheed, A. M. et al. The therapeutic potential of convalescent plasma therapy on treating critically-ill COVID-19 patients residing in respiratory care units in hospitals in Baghdad, Iraq. Infez. Med. 28, 357–366 (2020).

9. Agarwal, A. et al. Convalescent plasma in the management of moderate covid-19 in adults in India: open label phase II multicentre randomised controlled trial (PLACID Trial). bmj 371, (2020).

10. AlQahtani, M. et al. Randomized controlled trial of convalescent plasma therapy against standard therapy in patients with severe COVID-19 disease. medRxiv (2020).

11. Libster, R. et al. Early High-Titer Plasma Therapy to Prevent Severe Covid-19 in Older Adults. N. Engl. J. Med. 0, null.

12. Simonovich, V. A. et al. A Randomized Trial of Convalescent Plasma in Covid-19 Severe Pneumonia. N. Engl. J. Med. (2020).

13. Li, X. et al. Risk factors for severity and mortality in adult COVID-19 inpatients in Wuhan. J. Allergy Clin. Immunol. 146, 110–118 (2020).

14. Salazar, E. et al. Significantly Decreased Mortality in a Large Cohort of Coronavirus Disease 2019 (COVID-19) Patients Transfused Early with Convalescent Plasma Containing High-Titer Anti–Severe Acute Respiratory Syndrome Coronavirus 2 (SARS-CoV-2) Spike Protein IgG. Am. J. Pathol. (2020).

15. Joyner, M. J. et al. Early safety indicators of COVID-19 convalescent plasma in 5,000 patients. J. Clin. Invest. (2020).

16. Joyner, M. J. et al. Safety update: COVID-19 convalescent plasma in 20,000 hospitalized patients. Mayo Clin. Proc. (2020).

17. Joyner, M. J. et al. Effect of Convalescent Plasma on Mortality among Hospitalized Patients with COVID-19: Initial Three-Month Experience. medRxiv (2020) doi:10.1101/2020.08.12.20169359.

18. Lawther, P. J., Waller, R. E. & Henderson, M. Air pollution and exacerbations of bronchitis. Thorax 25, 525–539 (1970).

19. Waller, R. E. & Lawther, P. J. Some observations on London fog. Br. Med. J. 2, 1356 (1955).

20. Bhatraju, P. K. et al. Covid-19 in critically ill patients in the Seattle region—case series. N. Engl. J. Med. 382, 2012–2022 (2020).

21. Muggeo, V. M. & others. Segmented: an R package to fit regression models with broken-line relationships. R News 8, 20–25 (2008).

22. Muggeo, V. M. & Muggeo, M. V. M. Package ‘segmented’. Biometrika 58, 516 (2017).

23. Wagner, A. K., Soumerai, S. B., Zhang, F. & Ross-Degnan, D. Segmented regression analysis of interrupted time series studies in medication use research. J. Clin. Pharm. Ther. 27, 299–309 (2002).

24. Touloumis, A. R package multgee: A generalized estimating equations solver for multinomial responses. ArXiv Prepr. ArXiv:14105232 (2014).

25. Nooraee, N., Molenberghs, G. & van den Heuvel, E. R. Gee for longitudinal ordinal data: Comparing R-geepack, R-multgee, R-repolr, SAS-genmod, SPSS-genlin. Comput. Stat. Data Anal. 77, 70–83 (2014).

